# Application of Elastic Net Regression for Modeling COVID-19 Sociodemographic Risk Factors

**DOI:** 10.1101/2023.01.19.23284288

**Authors:** Tristan A. Moxley, Jennifer Johnson-Leung, Erich Seamon, Christopher Williams, Benjamin J. Ridenhour

**Affiliations:** Bioinformatics and Computational Biology Program, University of Idaho, Moscow, ID, USA; Department of Mathematics and Statistical Science, University of Idaho, Moscow, ID, USA; Institute for Modeling Collaboration and Innovation, Moscow, ID, USA

**Keywords:** COVID-19, Elastic net regression, Risk modeling, Sociodemographic factors

## Abstract

**Objectives:** COVID-19 has been at the forefront of global concern since its emergence in December of 2019. Determining the social factors that drive case incidence is paramount to mitigating disease spread. We gathered data from the Social Vulnerability Index (SVI) along with Democratic voting percentage to attempt to understand which county-level sociodemographic metrics had a significant correlation with case rate for COVID-19.

**Methods:** We used elastic net regression due to issues with variable collinearity and model overfitting. Our modelling framework included using the ten Health and Human Services regions as submodels for the two time periods 22 March 2020 to 15 June 2021 (prior to the Delta time period) and 15 June 2021 to 1 November 2021 (the Delta time period).

**Results:** Statistically, elastic net improved prediction when compared to multiple regression, as almost every HHS model consistently had a lower root mean square error (RMSE) and satisfactory *R*^2^ coefficients. These analyses show that the percentage of minorities, disabled individuals, individuals living in group quarters, and individuals who voted Democratic correlated significantly with COVID-19 attack rate as determined by Variable Importance Plots (VIPs).

**Conclusions:** The percentage of minorities per county correlated positively with cases in the earlier time period and negatively in the later time period, which complements previous research. In contrast, higher percentages of disabled individuals per county correlated negatively in the earlier time period. Counties with an above average percentage of group quarters experienced a high attack rate early which then diminished in significance after the primary vaccine rollout. Higher Democratic voting consistently correlated negatively with cases, coinciding with previous findings regarding a partisan divide in COVID-19 cases at the county level. Our findings can assist policymakers in distributing resources to more vulnerable counties in future pandemics based on SVI.

## 1. Introduction

Severe Acute Respiratory Syndrome Coronavirus 2 (SARS-CoV-2) has impacted the world since its emergence in December of 2019, with a global case total of approximately 631 million cases and global death total of approximately 6.6 million deaths as of November 2022 (The New York Times 2022). Effective measures to quell the pandemic and minimize hospitalizations and deaths have included quarantining, mask mandates, social distancing measures (Fazio et al. 2021), and vaccines (Centers for Disease Control and Prevention 2022). The COVID-19 pandemic is unusual in the typical epidemiological sense, as wealthier countries with better health infrastructure have higher reported attack rates than their less wealthy counterparts (Bollyky et al. 2022). The heightened attack rate in the United States is a cause for concern. The U.S. has the highest confirmed case count of any nation at 97.6 million cases (as of November 2022), more than doubling India (the second-highest total) (The New York Times 2022). Disparities in case reporting may influence this trend. However, it is still reasonable to expect that nations with more wealth and better access to prophylactic resources would have lower disease burden overall, given the effectiveness of vaccines and other preventative tools (Bates et al. 2022).

From the perspective of the host-agent-environment model (Tsui, Deng, and Pan 2020), social factors present as a natural explanation for some of these discrepancies. When considering social factors, it is important to make the distinction between intrinsic factors (i.e., factors that one cannot easily change; these may include race, ethnicity, socioeconomic status, etc.), and extrinsic factors (i.e., qualities and behaviors that one acquires or changes throughout their daily lives, such as political ideology, occupation, etc.). In cases where intrinsic factors correlate with cases, state or federal governments can allocate more resources to particular regions where more disadvantaged individuals may live based on pre-existing county-level risk evaluation metrics such as the Social Vulnerability Index (SVI) (Centers for Disease Control and Prevention 2022). In cases where extrinsic factors correlate with cases, state or federal governments can assist in improving infrastructure and protocols for at-risk demographics within more vulnerable regions, as well as informing the public of proper pandemic responses through reputable sources such as the Centers for Disease Control (CDC).

Intrinsic social factors are multi-factorial in their impact on COVID-19 spread (Hooper, Nápoles, and Pérez-Stable 2020). Studies have shown an adverse relationship between being a racial or ethnic minority (e.g., African American, Indigenous, Hispanic, etc.) and contracting COVID-19; this translates as higher incidences of severe cases and deaths in certain demographics (Gayle and Childress 2021). The increased disease burden on racial/ethnic minorities has several contributing factors. Some of the underlying trends stem from higher rates of comorbidity, living in more crowded living conditions (Hooper, Nápoles, and Pérez-Stable 2020), and decreased ability to social distance due to working lower-paying, “essential” jobs in retail, transportation, agriculture, etc. (Gayle and Childress 2021). Many of these disparities have been well-documented prior to COVID-19. African Americans are eight times more likely to contract HIV compared to Caucasians on average, yet coverage of pre-exposure prophylaxis for treating HIV is seven times higher in Caucasians than in African Americans (Harris et al. 2019). Given the history of healthcare disparity and comorbidity influencing epidemiological attack rate (Hooper, Nápoles, and Pérez-Stable 2020), it is important to identify and prioritize these groups for prophylactic resource allocation.

Political affiliation has been hypothesized to have affected the spread of COVID-19 in the U.S., as pandemic response rapidly became a highly-politicized in the spring of 2020. This politicization was accompanied by the rapid spread and endorsement of misinformation regarding the severity and origin of SARS-CoV-2 (Motta, Stecula, and Farhart 2020). At the state level, jurisdictions with Democrat leadership had more aggressive responses to COVID-19 on average (Grossman et al. 2020). Some states with Republican administrations—such as Idaho—were more socially lax with mask and vaccine mandates, and subsequently suffered high spikes in cases and deaths from Delta and Omicron (Baker 2022). Some ideological conservatives and religious fundamentalists “may see [scientific] experts as threatening to their social identities” (Merkley and Loewen 2021), and thus may respond to COVID-19 in a more lax and risk-prone manner. Furthermore, Trump’s higher-than-average approval among conservative citizens may have had additional effects (Bartels 2018). Conservative individuals who cited former president Donald Trump and his task force as their primary information outlet were far less likely to get vaccinated (Jurkowitz and Mitchell 2021). However, as vaccine hesitancy has historical connections to both extremes of the political spectrum, a more formal analysis is necessary to determine the extent to which political affiliation is a contributing factor towards COVID-19 spread or vaccine acceptance (Troiano and Nardi 2021).

Our research looks into the effect of the combination of social forces, both intrinsic and extrinsic, and demography on COVID-19 cases. The remainder of this paper is organized as follows: A brief mention of model development is provided, along with elaboration on data selection. Significant variables found through these analyses are shown and interpreted through the lens of pandemic response at the county level. Our results can inform pandemic resource allocation based on areas at higher social risk.

## 2. Methods

We analyze the relationship of several measures of social vulnerability along with 2020 presidential voting preference on the incidence of COVID-19 at the county level. Data on social measures were obtained from the CDC’s Social Vulnerability Index (SVI) (CDC/ATSDR 2018). SVI is a percentage-per-county metric which synthesizes 15 census variables into one vulnerability score. This index is typically used to allocate necessary resources to vulnerable counties during disaster responses. Democratic voting percentage from the 2020 presidential election (CNN Politics 2020) is used as a proxy for political ideology. The full set of explanatory data is given in Table 1. Our response variable is cumulative cases per 1000 individuals and was sourced from the New York Times (The New York Times 2022).

**Table 1.** Explanatory variable abbreviations and their descriptions for SVI and voting percentage

| Variable | Description |
| --- | --- |
| POV | Percentage of individuals below poverty estimate |
| UNEMP | Percentage of unemployed individuals |
| NOHSDP | Percentage of individuals with no high school diploma |
| AGE65 | Percentage of individuals over the age of 65 |
| AGE17 | Percentage of individuals under the age of 17 |
| DISABL | Percentage of individuals with a non-institutionalized disability |
| SNGPNT | Percentage of individuals who are single parents with a child below the age of 18 |
| MINRTY | Percentage of minorities (all persons except white, non-Hispanic) |
| LIMENG | Percentage of individuals over the age of 5 who speak English “less than well” |
| MUNIT | Percentage of housing in structures with 10 or more units |
| MOBILE | Percentage of mobile homes |
| CROWD | Percentage of occupied housing units with more people than rooms |
| NOVEH | Percentage of households with no vehicle available |
| GROUPQ | Percentage of persons in group quarters |
| pct | Percentage voting Democratic in 2020 Presidential Election |

Elastic net regression is chosen for this analysis due to its ability to mitigate both multicollinearity and model overfitting through regularization (Zou and Hastie 2005), as both of these issues surfaced in exploratory data analysis. All analyses were performed using R version 4.1.3 (R Core Team 2022). The underlying model equation is **Y** = **X***β* + *ϵ*. In the case of elastic net regression, the estimated regression coefficients *β* are determined by minimizing a penalized sum of squares. Specifically, the sum of squares is penalized by the *elastic net penalty* comprised of the *l*_1_ and squared *l*_2_ norm, and is dependent on hyperparameters *α* and *λ*. Here, *α* controls the *l*_1_*/l*_2_ mixing percentage, and *λ* alters the penalization weight. A more detailed explanation of the elastic net model is available in Appendix B. The **caret** package was employed for determining the optimal hyperparameters necessary for constructing the elastic net models (Figure A2 in Appendix A displays an example of this penalization process). Verification of these parameters is determined via 70/30 repeated cross-validation with 15 iterations.

We model COVID-19 case rates at the county level for the ten Department of Health and Human Services (HHS) regions and two time periods. The definition of the HHS regions is given in Table 2 (U.S. Department of Health and Human Services 2022). The first time period analyzed is 22 March 2020–15 June 2021, which we will refer to as “pre-Delta” hereafter. The second time period is 16 June 2021–1 November 2021, which we will refer to “Delta” hereafter. Thus we have 20 different models of case rates in the U.S. (10 regions × 2 time periods). For each model, we calculate the root mean squared error (RMSE) for both multiple regression (as a baseline) and elastic net regression to verify that elastic net is producing more robust models. In addition to RMSE, *R*^2^ coefficients for both elastic net and multiple regression are presented to demonstrate elastic net’s ability to mitigate model overfitting.

**Table 2.** HHS region numbers and their member states used in all analyses

| Number | States |
| --- | --- |
| 1 | CT, MA, ME, NH, RI, VT |
| 2 | NJ, NY |
| 3 | DE, MD, PA, VA, WV |
| 4 | AL, FL, GA, KY, MS, NC, SC, TN |
| 5 | IL, IN, MI, MN, OH, WI |
| 6 | AR, LA, NM, OK, TX |
| 7 | IA, KS, MO, NE |
| 8 | CO, MT, ND, SD, UT, WY |
| 9 | AZ, CA, NV |
| 10 | ID, OR, WA |

Variable Importance Plots (VIP) are used to determine coefficient significance due to elastic net regression’s lack of theoretically derived *p*-values. Importance is calculated based on the absolute value of the coefficients (Greenwell and Boehmke 2020). Plots include both the individual variable importance values for each HHS region per time period, as well as box plots to view the average importance of each explanatory variable. From the full set of 15 explanatory variables, the five variables which exhibited the most noteworthy correlations with cases are isolated and interpreted. Appendix C contains the full models with all 15 explanatory variables. Plots of the observed data and the associated predicted regression line for each region are also provided to assess individual model performance.

## 3. Results

All coefficient estimates are reported in their original scale, allowing for direct interpretation. Overall, the RMSEs for elastic net models of the test data sets were lower than their multiple regression counterparts in all but two of the regions (HHS region 5 for pre-Delta, HHS region 6 for Delta); this reduction in RMSE indicates more robust model performance for the elastic net models. Individual model attributes will be discussed in their respective subsections.

The pre-Delta coefficient estimates and metrics are shown in Table 3. In regions 3,6, and 10, elastic net regression removes several features from the regression model (see Table C2). The small difference in training/testing *R*^2^ for elastic net regression (Table 3) demonstrates that this method of regression avoids the problem of model overfitting. Figure 1 displays the VIP plot for the pre-Delta and Delta COVID-19 time periods. As shown in Figure 1a, we find that the percentage of individuals living in group quarters has the highest overall variable importance across all regions, with Democratic voting percentage and percentage of individuals in mobile homes being the other most important variables, respectively (Figure 1a). Of the selected variables, percentage non-institutionalized disabled individuals is least significant.

**Table 3.** Coefficients and metrics for the 10 HHS regions for the pre-Delta COVID-19 time period, recorded from March 22, 2020 to June 15, 2021

| Coefficients |  |  |  |  |  |  |  |  |  |  |
| --- | --- | --- | --- | --- | --- | --- | --- | --- | --- | --- |
| Region | 1 | 2 | 3 | 4 | 5 | 6 | 7 | 8 | 9 | 10 |
| (Intercept) | 65.14 | 87.20 | 85.06 | 109.20 | 106.13 | 107.57 | 106.38 | 113.58 | 88.21 | 71.52 |
| Disability | -2.95 | - | - | - | 3.21 | - | -0.84 | -4.60 | -6.11 | - |
| Minority | 3.71 | 4.59 | - | 5.07 | 8.18 | - | -1.13 | - | 24.01 | - |
| Mobile Housing | -12.5 | -7.95 | - | -6.16 | -3.67 | -2.85 | -6.61 | -7.19 | - | -1.03 |
| Group Quarters | -0.59 | -3.61 | 11.00 | 6.66 | 5.29 | 6.43 | 6.21 | 17.06 | 13.59 | 4.11 |
| Voting Percentage | -5.68 | -3.99 | -8.33 | -9.38 | -10.50 | -0.87 | -1.07 | -1.77 | -11.21 | -9.85 |
| Metrics |  |  |  |  |  |  |  |  |  |  |
| $\alpha$ | 0.10 | 0.19 | 0.50 | 0.91 | 0.36 | 0.54 | 0.73 | 0.81 | 0.27 | 0.42 |
| $\lambda$ | 4.31 | 2.61 | 0.46 | 0.20 | 0.09 | 2.10 | 0.37 | 0.99 | 0.81 | 3.58 |
| ENR Train $R^2$ | 0.78 | 0.78 | 0.44 | 0.20 | 0.41 | 0.28 | 0.35 | 0.40 | 0.71 | 0.66 |
| ENR Test $R^2$ | 0.78 | 0.78 | 0.42 | 0.15 | 0.37 | 0.24 | 0.35 | 0.37 | 0.64 | 0.64 |
| MR Train $R^2$ | 0.82 | 0.84 | 0.50 | 0.22 | 0.39 | 0.30 | 0.38 | 0.48 | 0.73 | 0.75 |
| MR Test $R^2$ | 0.76 | 0.63 | 0.34 | 0.11 | 0.32 | 0.20 | 0.30 | 0.25 | 0.46 | 0.60 |
| ENR Test RMSE | 15.48 | 11.52 | 12.88 | 20.00 | 15.68 | 23.95 | 21.05 | 28.11 | 26.79 | 19.27 |
| MR Test RMSE | 17.49 | 16.66 | 13.12 | 20.09 | 15.67 | 33.60 | 21.10 | 29.25 | 28.43 | 19.52 |

**Figure 1.**
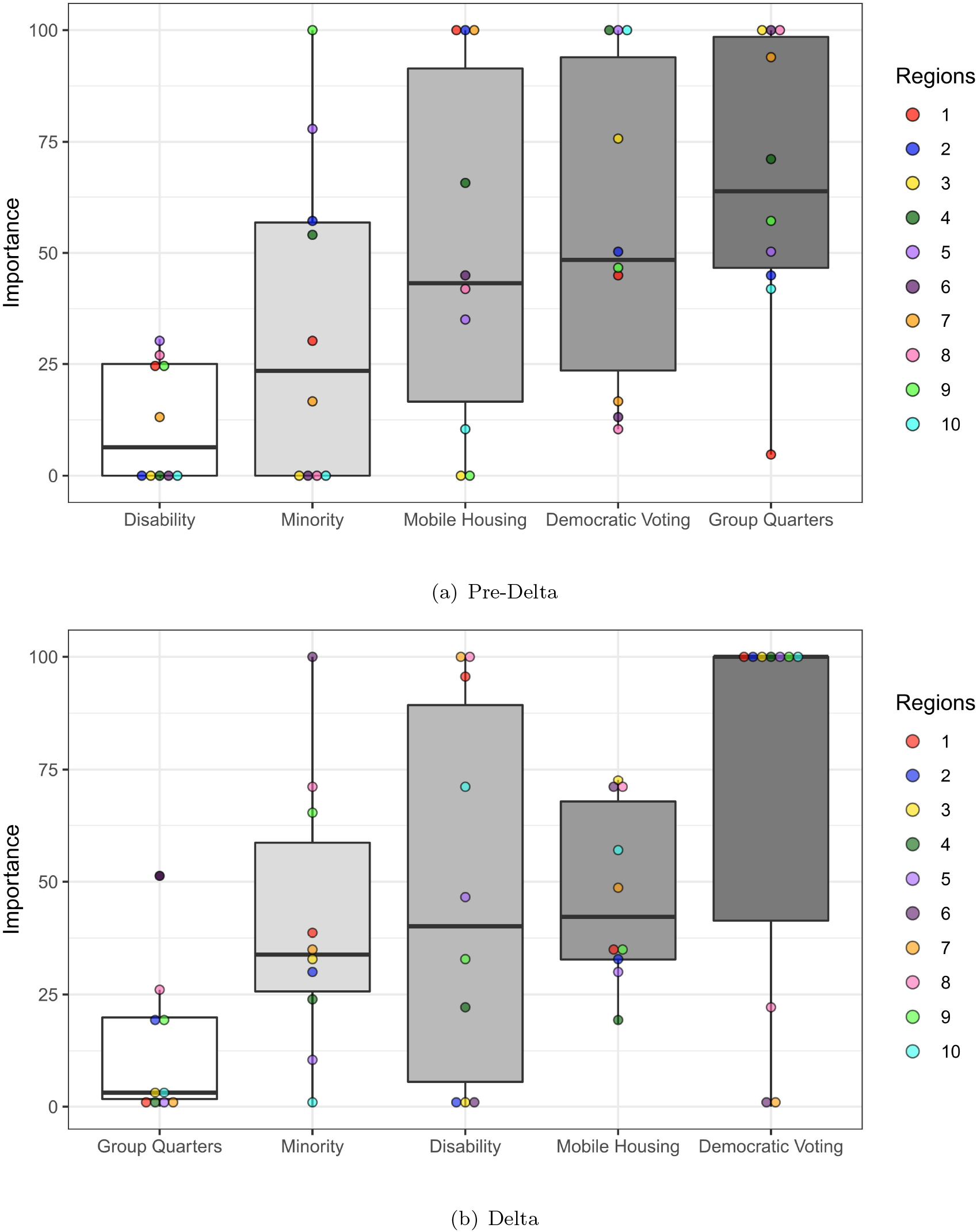
Variable Importance Plots for both Pre-Delta and Delta time periods, organized from lowest to highest overall importance

The Delta coefficient estimates and metrics are shown in Table 4. While there is some change in the features selected by the elastic net regression models in this time period, the *R*^2^ comparisons for the testing and training data sets show that the method effectively avoids the problem of overfitting. Figure 1b displays the VIP plots for the Delta time period. The variable of lowest importance in the Delta time period is the percentage of individuals living in group quarters. Democratic voting percentage has the highest variable importance across all regions, followed by the percentage of households living in mobile homes and the percentage of disabled individuals per county. Figure 1b shows that voting percentage has the highest individual variable importance in seven of the ten HHS regions for the Delta time period. Of the selected variables, group quarters has the largest shift in importance, going from most important to least important, whereas disability percentage increases dramatically in significance from pre-Delta to Delta.

**Table 4.** Coefficients and metrics for the 10 HHS regions for the Delta COVID-19 time period, recorded from June 15, 2021 to November 1, 2021

| Coefficients |  |  |  |  |  |  |  |  |  |  |
| --- | --- | --- | --- | --- | --- | --- | --- | --- | --- | --- |
| Region | 1 | 2 | 3 | 4 | 5 | 6 | 7 | 8 | 9 | 10 |
| (Intercept) | 23.50 | 24.60 | 49.69 | 61.08 | 40.32 | 49.07 | 36.03 | 39.92 | 40.47 | 46.76 |
| Disability | 1.31 | – | – | 2.20 | 3.77 | – | 3.76 | 6.64 | 2.57 | 4.16 |
| Minority | –0.53 | –0.69 | –2.30 | –2.34 | 0.85 | –1.91 | –1.34 | –4.76 | –4.99 | – |
| Mobile Housing | 0.49 | –0.72 | 5.21 | 1.81 | 2.35 | 1.37 | 1.83 | 4.74 | –2.61 | 3.37 |
| Group Quarters | – | –0.45 | –0.29 | 0.19 | 0.13 | –0.98 | – | –1.73 | –1.46 | 0.14 |
| Voting Percentage | –1.37 | –2.23 | –7.18 | –9.78 | –8.09 | – | – | –1.45 | –7.64 | –5.91 |
| Metrics |  |  |  |  |  |  |  |  |  |  |
| $\alpha$ | 0.30 | 0.84 | 0.71 | 0.66 | 0.67 | 0.90 | 0.95 | 0.12 | 0.14 | 0.20 |
| $\lambda$ | 1.94 | 0.19 | 1.76 | 0.14 | 0.07 | 0.78 | 0.60 | 0.98 | 0.98 | 1.18 |
| ENR Train $R^2$ | 0.47 | 0.62 | 0.72 | 0.46 | 0.55 | 0.16 | 0.24 | 0.28 | 0.73 | 0.60 |
| ENR Test $R^2$ | 0.46 | 0.55 | 0.72 | 0.41 | 0.51 | 0.16 | 0.27 | 0.25 | 0.69 | 0.59 |
| MR Train $R^2$ | 0.61 | 0.72 | 0.84 | 0.47 | 0.57 | 0.21 | 0.31 | 0.30 | 0.76 | 0.70 |
| MR Test $R^2$ | 0.15 | 0.29 | 0.57 | 0.37 | 0.45 | 0.07 | 0.22 | 0.18 | 0.62 | 0.46 |
| ENR Test RMSE | 7.04 | 4.49 | 12.33 | 13.09 | 9.15 | 15.21 | 9.96 | 15.05 | 9.30 | 14.09 |
| MR Test RMSE | 12.05 | 4.77 | 14.83 | 13.16 | 9.18 | 14.92 | 10.36 | 16.04 | 9.79 | 14.86 |

Figure 2 displays the model fit for both pandemic time periods across each region, with each individual point having opacity and size based on population density (e.g., a county with low population density will have a small, transparent data point, and vice versa for a county with high population density). Regions 1, 2, 3, 9, and 10 (i.e., regions along the coast) present much better overall model fit when compared to the other five HHS regions (i.e., the inland regions), as shown through each model’s *R*^2^ coefficient. Overall, most models fit the data well, being able to explain a fair proportion of the variation within noisy case data.

**Figure 2.**
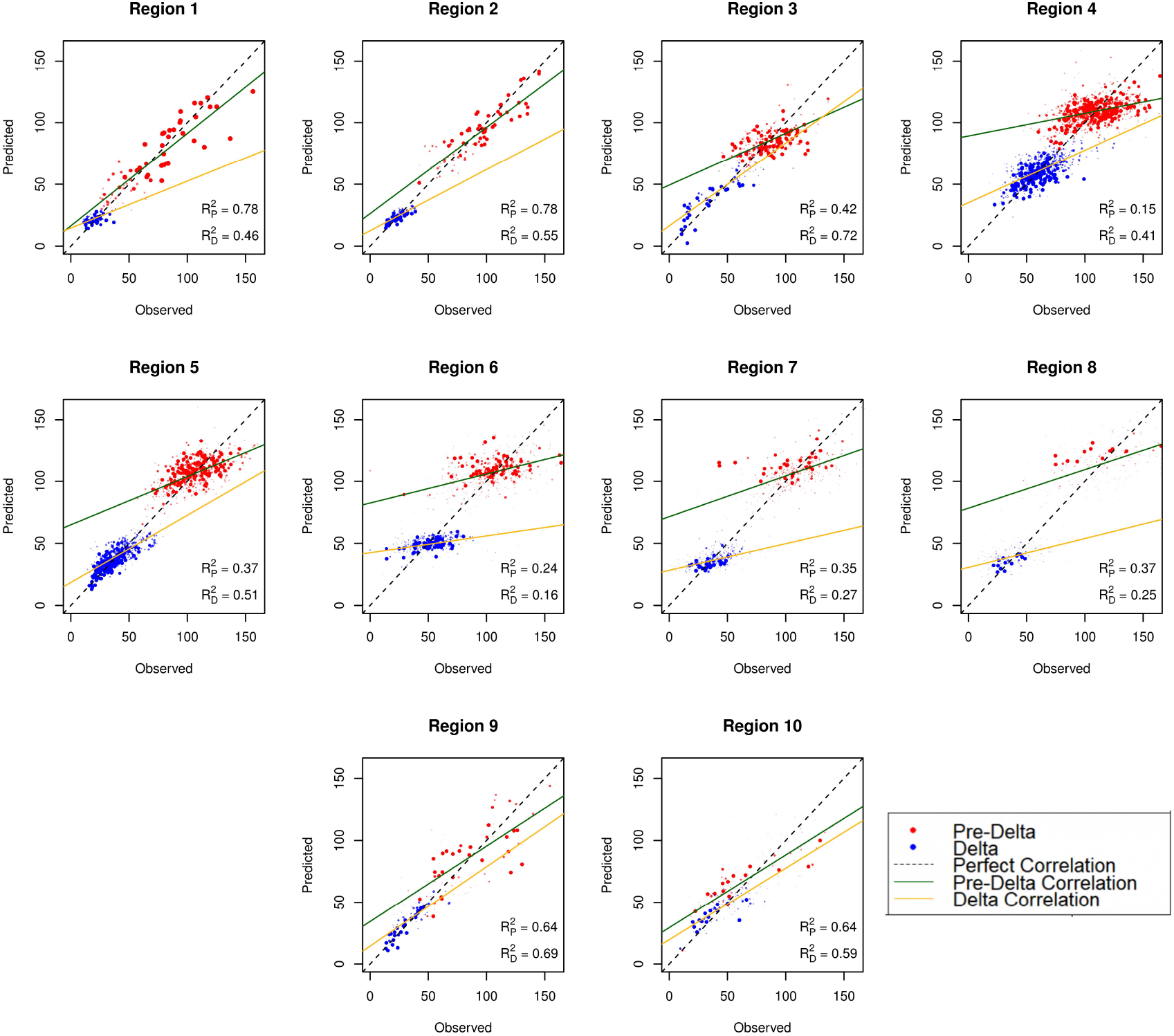
Observed versus predicted COVID-19 case rate plot for both pandemic time periods, used to visualize model performance. The “perfect” correlation line shows a perfect one-to-one relationship between the observed data and the fitted data, whereas each time period-specific line shows the observed correlation for each respective pandemic time period.

## 4. Discussion

Elastic net regression improves the prediction of COVID-19 cases when compared to multiple regression. All but two of the HHS regions across both the pre-Delta and Delta time periods have a lower testing set RMSE when compared to their multiple regression counterparts. Model fit and *R*^2^ coefficients vary based on region, with inland regions having lower *R*^2^ values. The regions with lower *R*^2^ values also happen to be ones with more counties of lower population density. Regions 1, 2, 3, 9, and 10 have a combined average population density of about 467 people per square mile, whereas Regions 4, 5, 6, 7, and 8 have a combined population density of about 115 people per square mile. This implies that, using our methods, relationships between the social vulnerability measures and disease burden are more difficult to observe in regions with lower population density. This is important to consider when planning for future mitigation of health emergencies.

It was known early on in the pandemic that close-quarters such as cruise ships were epicenters of COVID-19 spread (Sloane 2020). Similarly, increasing the county-wide percentage of people living in group quarters, such as nursing homes and prisons, has also been shown to increase risk (Karmakar, Lantz, and Tipirneni 2021; Millar et al. 2021). Our results show that this continued to be important through the pre-Delta time period of the pandemic. For the pre-Delta time period, the percentage of individuals in group quarters has the highest average variable importance across all regions. In all but regions 1 and 2, there is a strong positive association with cases. However, group quarters has the lowest average variable importance for the Delta time period. There are several possible explanations for this. Vaccination roll-out early on in 2021 was shown to mitigate disease spread (White et al. 2021). In many locales, scarce vaccination doses were prioritized for these populations and vaccine acceptance among the elderly and institutionalized was relatively high (Centers for Disease Control and Prevention 2022). Additionally, the lower association between the percentage of residents living in group quarters and COVID-19 cases during the Delta time period could be explained by a larger initial attack rate within these counties. Indeed, many prisons were severely unequipped to mitigate COVID-19 transmission and thus suffered mass infections during the pre-Delta time period (Burki 2020).

The percentage of non-institutionalized disabled persons per county was one of the least important variables in the pre-Delta time period and one of the most important variables in the Delta time period. The spectrum of what is considered disability is quite broad and encompasses many conditions (e.g., impairment of hearing, sight, mobility, or cognition) and thus the percentage of disabled people can be quite high (15.4%). Because the definition of disability by the American Community Survey (ACS) is broad, it makes our findings with respect to the percentage of disabled persons harder to interpret. The pre-Delta finding indicates the percentage of disabled people did not correlate with overall case counts. This is somewhat surprising; not only were many governments devoid of any resources for disabled individuals in their pandemic responses, but several factors put disabled individuals at an increased risk for infection, such as dependence on caretakers creating often unavoidable exposure (Armitage and Nellums 2020) and non-accessible hygiene and informational resources for individuals with visual, auditory, and cognitive disabilities (Kavanagh et al. 2022). However, the lack of relationship could either be due to the broad classification being used, or it could be that the fraction of individuals actually requiring specialized care is low enough that it did not affect the overall spread of COVID-19. We did observe the aforementioned trend of percentage of disabled persons predicting case counts in the Delta time period. It unclear why this would become a significant predictor later in the pandemic, however, it may be due to increased population mixing/reduced social distancing that occurred during this time period. If reduced social distancing is the cause, it might reflect that this population generally remained healthy early and thus acted as an influx of susceptible individuals later. It should be pointed out that looking at COVID-19 related deaths might have stronger relationship with the size of the disabled population, as the underlying comorbidities for disabled individuals present a higher likelihood of increased deaths rather than cases (Turk et al. 2020).

Percentage of minorities per-county was important across both pandemic time periods, with slightly higher importance in the Delta time period when transmissibility increased and mandated quarantining decreased. For the pre-Delta time period, all coefficients held positive values, which coincides with previous county-level research citing positive correlations with COVID-19 cases and minority proportions per county (Lee et al. 2021). This is complementary to previous research citing a disproportionate impact of COVID-19 on minority individuals due to heightened levels of comorbidity, general inability to distance, and worse living conditions on average (Gayle and Childress 2021). However, the trend reverses in the Delta time period, with many of the HHS regions having negative coefficient values. Like with group quarters, this could be due to a disproportionately high attack rate during the first major wave of the pandemic. Much like with disabled individuals, an analysis that takes into account COVID-19 deaths and hospitalizations may give more credence to the disproportionate toll of COVID-19 on counties with a higher proportion of minorities, given that many comorbidities minorities experience result in more severe infections regardless of SARS-CoV-2 variant (Hooper, Nápoles, and Pérez-Stable 2020).

Democratic voting percentage had highly significant negative correlations with cases across both pandemic time periods. This is supported by previous studies which cite a county-level partisan correlation (as determined by voting percentage) with respect to COVID-19 distancing (Roberts and Utych 2021), cases (Gollwitzer et al. 2020), and deaths (Wallace, Goldsmith-Pinkham, and Schwartz 2022). Additionally, these findings complement previous research which cites a partisan divide with respect to trust in pandemic information resources and the effect it had on COVID-19 attack rates (Latkin et al. 2020). Given that vaccines were highly effective in reducing case incidence up through the Delta variant (Bernal et al. 2021) and that Republican individuals reportedly had 90% lower odds of vaccination when compared to Democrats (Dolman et al. 2022), disease behavior along partisan lines appears to have a significant effect on disease spread at the county level. Presidential voting is not the most foolproof proxy for the overall effect of political leaning; it fails to capture much of the extrinsic social effects caused by such factors as local legislature, personal risk evaluation, and media influence. However, it is still useful for potential policy inferences due to its availability and census-level information.

### Limitations

Although our methods and data were carefully chosen, some limitations still exist. The analysis conducted on the pre-Delta time period had a much larger pool of data when compared to the Delta model, since the pre-Delta model contained 15 months worth of cumulative COVID-19 data, whereas the Delta model only contained five months. Further subdivision of COVID-19 time periods may be an appropriate approach for refining this research by separating the models out into the original strain, Alpha, Delta, Omicron, etc. Additionally, many social factors were not considered in this research. As alluded to when discussing political influences, vaccine hesitancy is a highly-important extrinsic social factor that has clear implications on local-level COVID-19 attack rates. Inclusion of vaccine hesitancy (or, at minimum, a proxy for this factor), would likely yield significant results. Another potential confounding variable is population density; model fit discrepancies according to population density (Figure 2) may imply population density as an influential variable in spite of response variable standardization. A more robust analysis using our modeling techniques would include more social factors of perceived importance as well as confounders such as population density.

We chose to use case rates (i.e., case count per 1000 people) as our response variable for the elastic net regression models. Standardizing the response variables in this method allows for use of standard multiple regression along with elastic net regression, as the case rates approximately follow a normal distribution (Figure A1). Previous analyses by Karmakar, Lantz, and Tipirneni and Millar et al., use similarly scaled COVID-19 cases/deaths (with Karmakar, Lantz, and Tipirneni (2021) using cases per 100,000 and Millar et al. (2021) using adjusted case-fatality rate). However, both of these analyses opt for using negative binomial regression via generalized linear models as the response variable is count data at its core. Thus, this analysis takes a different modeling approach when compared to other prior analyses. The net effect of choosing to use a normally distributed error versus some other model is a shift in parameter confidence intervals. Because we use variable importance to judge the reliability of our predictors, confidence intervals of parameters are not relevant.

Finally, we believe that these analyses may benefit from using deaths or hospitalizations as the response variable, rather than case counts. As seen with group quarters and minorities per county, the overall course of case spread does not necessarily display the broader picture of disease severity according to the pre-existing research (Sloane 2020; Gayle and Childress 2021). Cases can only predict so much; vaccine and natural immunity can suppress the perceived effect on individuals who are either at-risk of infection due to their environment, or are at-risk of severe illness due to autoimmune disorders or comorbidities. The application of this type of method to hospitalizations and deaths can help to enhance these results. In general, more consideration in using additional nuanced predictors along with repeating the analyses with hospitalizations and/or deaths would be valuable when thinking about future disease response.

## 5. Conclusion

Understanding the relationship between COVID-19 case rates and existing measures of social vulnerability can help inform future pandemic planning. We found a mixture of intrinsic (disability and minority percentages in counties) and extrinsic (Democrat voting, mobile housing, and group quarters percentages) social factors. Policy decisions for emerging pandemics have two avenues: resource allocation (e.g., vaccines, masks, etc.) to areas that have higher attack rates due to intrinsic factors, and implementing measures (e.g., information campaigns) prior to disease outbreaks to better prepare vulnerable areas with higher extrinsic risk factors. In other words, our results suggest physical pandemic resources should be allocated toward counties with higher social vulnerability measures such as those with larger proportions of minorities or disabled individuals. Conversely, it has been observed that the political climate within the United States diminished the trustworthiness of many public information outlets like CDC (Latkin et al. 2020). Our findings, as well as those of others, suggest an appropriate prophylactic measure would be establishing trusted information sources in preparation for future outbreaks. Providing a unified, trusted, non-partisan outlet for pandemic information can ensure the public receives accurate messages regarding disease prevention. Future research may involve either expanding the feature set provided in the analysis to better understand the county-level correlations present in COVID-19 case data, or in looking more deeply into the significant variables found within this analysis to better understand the correlations.

## Data Availability Statement

The data that support the findings of this study are available in Figshare at 10.6084/m9.figshare.21777674. These data were derived from the following resources available in the public domain: CDC/ATSDR (https://www.atsdr.cdc.gov/placeandhealth/svi/data_documentation_download.html), New York Times (https://github.com/nytimes/covid-19-data), and CNN (https://www.cnn.com/election/2020/results/president).

### Appendix A. Supplementary Figures

**Figure A1.**
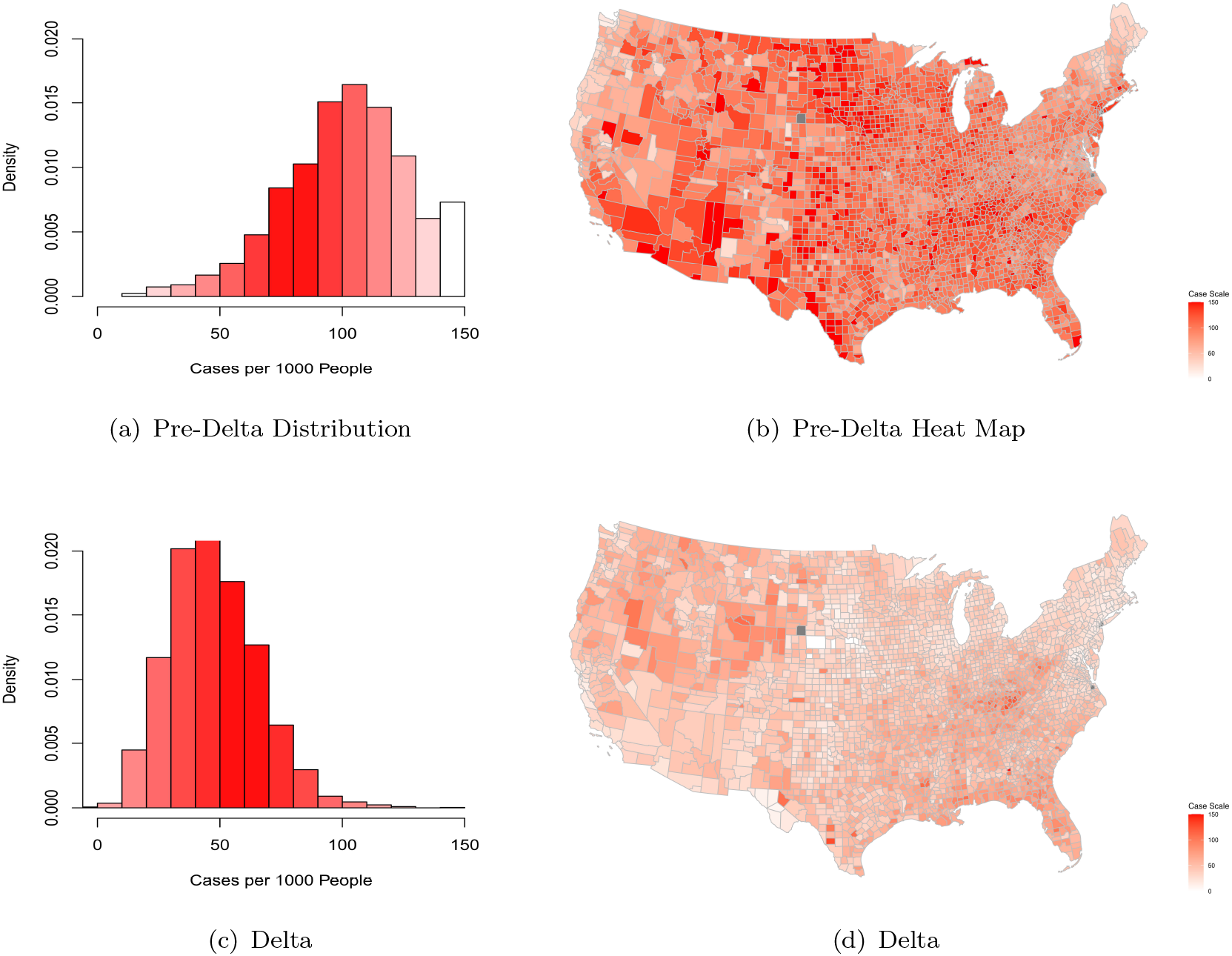
Case probability distributions and heat maps for both pre-Delta and Delta pandemic time periods

**Figure A2.**
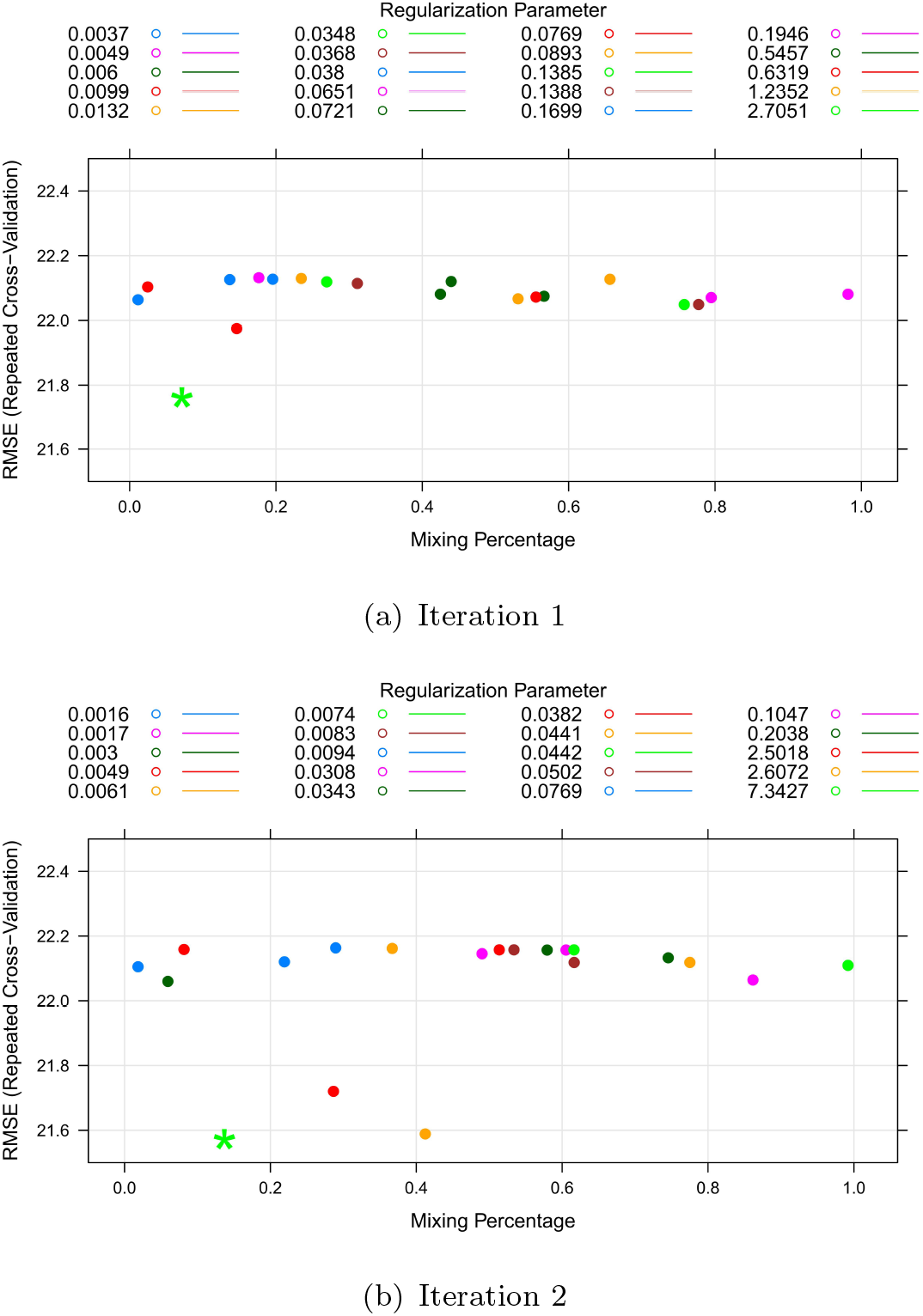
Display of varying optimal hyperparameters across two iterations of generating the elastic net model for the Pacific Northwest HHS region

### Appendix B. Elastic Net Regression Overview

Elastic net regression is a regularization technique developed by Hui Zou and Trevor Hastie to overcome the limitations of *l*_1_ (lasso) and *l*_2_ (ridge) regularization (Zou and Hastie 2005). *l*_1_ regularization is a penalty to the OLS estimators that results in feature selection, selecting against unimportant variables, whereas *l*_2_ regularization is a penalty to the OLS (Ordinary Least Squares) estimators that shrinks correlated predictors towards each other to overcome multicollinearity (Zou and Hastie 2005). What elastic net achieves is a balance between these two penalties, resulting in a process by which models can be created that deal with multicollinearity among explanatory variables while simultaneously selecting the important features out of a large set of potential predictors. Zou and Hastie (paired with simplified derivations from Friedman, Hastie, and Tibshirani (2010)) describe the set up for the model as follows:

Suppose we have a vector **Y** = (*y*_1_, …, *y*_*n*_), which is our observed data, and **X** = (*x*_1_| … |*x*_*k*_) be our model matrix. Also suppose we have **x**_*j*_ = (*x*_1*j*_, …, *x*_*nj*_)^*T*^, *j* = 1, …, *p* are the predictor variables. We can construct a regression model such that **Y** = *β*_0_ + *x*^*T*^ *β* + *ϵ*, where *β*_0_ is the intercept of the regression equation, *β* are the coefficients for the *p* explanatory variables, and *ϵ* is our residual vector. For the elastic net penalty, we assume that the response is centered and the predictors are standardized. This means that the sum of our observed data should equal 0, the sum of our explanatory variables should equal 0, and the mean squared error of our explanatory variables should equal 1. This can be represented as:

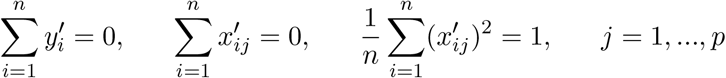

Here, 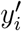 is the centered observed response for the *i*^*th*^ county, and 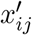 is the standardized measurement for the *j*^*th*^ explanatory variable in the *i*^*th*^ county. Once this pre-processing has been accomplished, we define the model. The proposition for this model is similar to that of a standard regression model. In standard OLS, we wish to minimize the sum of squared residuals in order to fit a linear model to the data. This is similar to that approach, but with the added *l*_1_ and *l*_2_ penalties. As such, we wish to find:

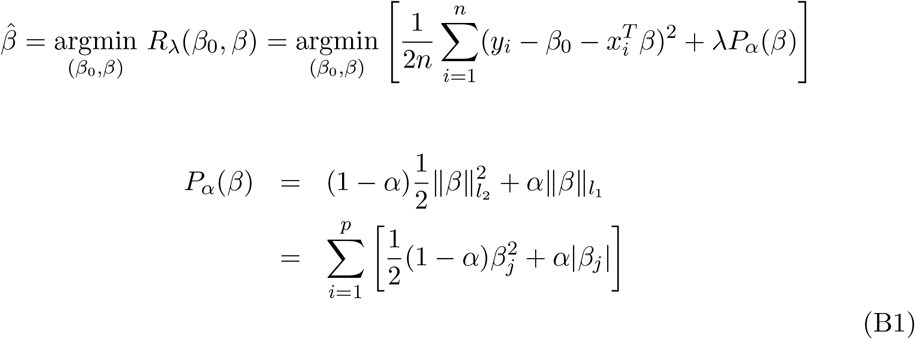

*α* and *λ* are the hyperparameters for this model, and help to determine the mixture and weight of penalization, respectively. Intuitively, we can see why elastic net is a generalization of lasso and ridge. Setting *α* = 1 results in the elastic net penalty only contributing a penalty equivalent to the magnitude of the *β* coefficients (also known as the *l*_1_ norm), thus resulting in strictly *l*_1_ regularization. Conversely, setting *α* = 0 results in the elastic net penalty only contributing a penalty equivalent to the squared Euclidean distance of the *β* coefficients (also known as the *l*_2_ norm), thus resulting in strictly *l*_2_ regularization. These penalties together are scaled by *λ*; the higher the value for *λ*, the higher the penalty accrued to the regression.

### Appendix C. Full Model Tables and VIPs

**Table C1.** Coefficients and metrics for the 10 HHS regions for the pre-Delta COVID-19 time period, recorded from March 22, 2020 to June 15, 2021

| Coefficients |  |  |  |  |  |  |  |  |  |  |
| --- | --- | --- | --- | --- | --- | --- | --- | --- | --- | --- |
| Region | 1 | 2 | 3 | 4 | 5 | 6 | 7 | 8 | 9 | 10 |
| (Intercept) | 65.14 | 87.20 | 85.06 | 109.20 | 106.13 | 107.57 | 106.38 | 113.58 | 88.21 | 71.52 |
| In Poverty | -2.46 | - | -0.77 | - | -1.43 | - | -5.51 | -1.80 | 10.72 | 3.16 |
| Unemployed | -0.14 | 0.40 | -1.28 | -1.19 | -5.22 | - | - | 1.27 | 1.54 | -1.07 |
| No HS Diploma | 3.70 | - | -1.66 | 3.82 | -2.63 | 2.17 | 3.84 | 6.36 | 2.35 | - |
| Over 65 | -3.63 | - | -0.97 | -1.89 | - | -1.91 | 2.92 | - | 8.18 | -6.14 |
| Under 17 | 2.30 | 5.55 | 0.65 | 2.50 | 1.77 | 4.55 | 1.67 | 2.24 | 4.27 | 12.32 |
| Disability | -2.95 | - | - | - | 3.21 | - | -0.84 | -4.60 | -6.11 | - |
| Single Parent | 2.17 | -1.42 | 2.78 | - | 3.33 | 4.19 | 6.68 | 8.36 | 0.84 | - |
| Minority | 3.71 | 4.59 | 0 | 5.07 | 8.18 | - | -1.13 | - | 24.01 | - |
| Limited English | 6.87 | 6.20 | 4.60 | 0.31 | 3.53 | - | 5.19 | -4.76 | - | - |
| Multi-Unit Home | - | 0.56 | -0.14 | 0.36 | 2.02 | - | 1.73 | 3.96 | 4.40 | 2.59 |
| Mobile Housing | -12.45 | -7.95 | 0 | -6.16 | -3.67 | -2.85 | -6.61 | -7.19 | - | -1.03 |
| Crowded Housing | -2.00 | 2.47 | 1.05 | 1.08 | - | 0.76 | 3.01 | 1.80 | -3.81 | - |
| No Vehicle | - | -0.89 | 3.58 | - | -3.55 | 1.06 | -0.19 | - | -4.65 | -1.85 |
| Group Quarters | -0.59 | -3.61 | 11.00 | 6.66 | 5.29 | 6.43 | 6.21 | 17.06 | 13.59 | 4.11 |
| Voting Percentage | -5.68 | -3.99 | -8.33 | -9.38 | -10.50 | -0.87 | -1.07 | -1.77 | -11.21 | -9.85 |
| Metrics |  |  |  |  |  |  |  |  |  |  |
| $\alpha$ | 0.10 | 0.19 | 0.5 | 0.91 | 0.36 | 0.54 | 0.73 | 0.81 | 0.27 | 0.42 |
| $\lambda$ | 4.31 | 2.61 | 0.46 | 0.20 | 0.09 | 2.10 | 0.37 | 0.99 | 0.81 | 3.58 |
| ENR Train $R^2$ | 0.78 | 0.78 | 0.44 | 0.20 | 0.41 | 0.28 | 0.35 | 0.40 | 0.71 | 0.66 |
| ENR Test $R^2$ | 0.78 | 0.78 | 0.42 | 0.15 | 0.37 | 0.24 | 0.35 | 0.37 | 0.64 | 0.64 |
| MR Train $R^2$ | 0.82 | 0.84 | 0.50 | 0.22 | 0.39 | 0.30 | 0.38 | 0.48 | 0.73 | 0.75 |
| MR Test $R^2$ | 0.76 | 0.63 | 0.34 | 0.11 | 0.32 | 0.20 | 0.30 | 0.25 | 0.46 | 0.60 |
| ENR Test RMSE | 15.48 | 11.52 | 12.88 | 20.00 | 15.68 | 23.95 | 21.05 | 28.11 | 26.79 | 19.27 |
| MR Test RMSE | 17.49 | 16.66 | 13.12 | 20.09 | 15.67 | 33.60 | 21.10 | 29.25 | 28.43 | 19.52 |

**Table C2.** Coefficients and metrics for the 10 HHS regions for the Delta COVID-19 time period, recorded from June 15, 2021 to November 1, 2021

| Coefficients |  |  |  |  |  |  |  |  |  |  |
| --- | --- | --- | --- | --- | --- | --- | --- | --- | --- | --- |
| Region | 1 | 2 | 3 | 4 | 5 | 6 | 7 | 8 | 9 | 10 |
| (Intercept) | 23.50 | 24.60 | 49.69 | 61.08 | 40.32 | 49.07 | 36.03 | 39.92 | 40.47 | 46.76 |
| In Poverty | 1.38 | 1.25 | 5.15 | 2.82 | 1.17 | 0.31 | - | -2.51 | 2.69 | -0.30 |
| Unemployed | - | 1.26 | - | 1.01 | -2.09 | - | 1.01 | 2.72 | -1.23 | 2.53 |
| No HS Diploma | 0.23 | - | - | -1.19 | -1.96 | 0.24 | - | -3.69 | 0.19 | -0.20 |
| Over 65 | - | - | - | 0.76 | -0.52 | -2.49 | -1.26 | -4.95 | -7.41 | -1.68 |
| Under 17 | - | 0.26 | - | 3.73 | 1.44 | - | - | -0.44 | 3.04 | 2.21 |
| Disability | 1.31 | - | - | 2.20 | 3.77 | - | 3.76 | 6.64 | 2.57 | 4.16 |
| Single Parent | 0.52 | 1.00 | - | 1.12 | 1.87 | 0.90 | 1.48 | -0.40 | 0.23 | 4.70 |
| Minority | -0.53 | -0.69 | -2.30 | -2.34 | 0.85 | -1.91 | -1.34 | -4.76 | -4.99 | - |
| Limited English | - | -2.03 | - | - | -0.44 | -3.23 | - | 0.02 | -2.23 | -0.90 |
| Multi-Unit Home | -1.03 | - | - | 4.58 | 2.36 | - | - | 0.95 | -4.62 | - |
| Mobile Housing | 0.49 | -0.72 | 5.21 | 1.81 | 2.35 | 1.37 | 1.83 | 4.74 | -2.61 | 3.37 |
| Crowded Housing | - | - | - | 0.88 | -0.74 | 0.12 | - | 2.20 | -1.56 | -2.33 |
| No Vehicle | - | - | - | -0.09 | 0.43 | 1.65 | 0.17 | 2.17 | 3.61 | 2.35 |
| Group Quarters | - | -0.45 | -0.29 | 0.19 | 0.133 | -0.98 | - | -1.73 | -1.46 | 0.14 |
| Voting Percentage | -1.37 | -2.23 | -7.18 | -9.78 | -8.09 | - | - | -1.45 | -7.64 | -5.91 |
| Metrics |  |  |  |  |  |  |  |  |  |  |
| $\alpha$ | 0.30 | 0.84 | 0.71 | 0.66 | 0.67 | 0.90 | 0.95 | 0.12 | 0.14 | 0.20 |
| $\lambda$ | 1.94 | 0.19 | 1.76 | 0.14 | 0.07 | 0.78 | 0.60 | 0.98 | 0.98 | 1.18 |
| ENR Train $R^2$ | 0.47 | 0.62 | 0.72 | 0.46 | 0.55 | 0.16 | 0.24 | 0.28 | 0.73 | 0.60 |
| ENR Test $R^2$ | 0.46 | 0.55 | 0.72 | 0.41 | 0.51 | 0.16 | 0.27 | 0.25 | 0.69 | 0.59 |
| MR Train $R^2$ | 0.61 | 0.72 | 0.84 | 0.47 | 0.57 | 0.21 | 0.31 | 0.30 | 0.76 | 0.70 |
| MR Test $R^2$ | 0.15 | 0.29 | 0.57 | 0.37 | 0.45 | 0.07 | 0.22 | 0.18 | 0.62 | 0.46 |
| ENR Test RMSE | 7.04 | 4.49 | 12.33 | 13.09 | 9.15 | 15.21 | 9.96 | 15.05 | 9.30 | 14.09 |
| MR Test RMSE | 12.05 | 4.77 | 14.83 | 13.16 | 9.18 | 14.92 | 10.36 | 16.04 | 9.79 | 14.86 |

**Figure C1.**
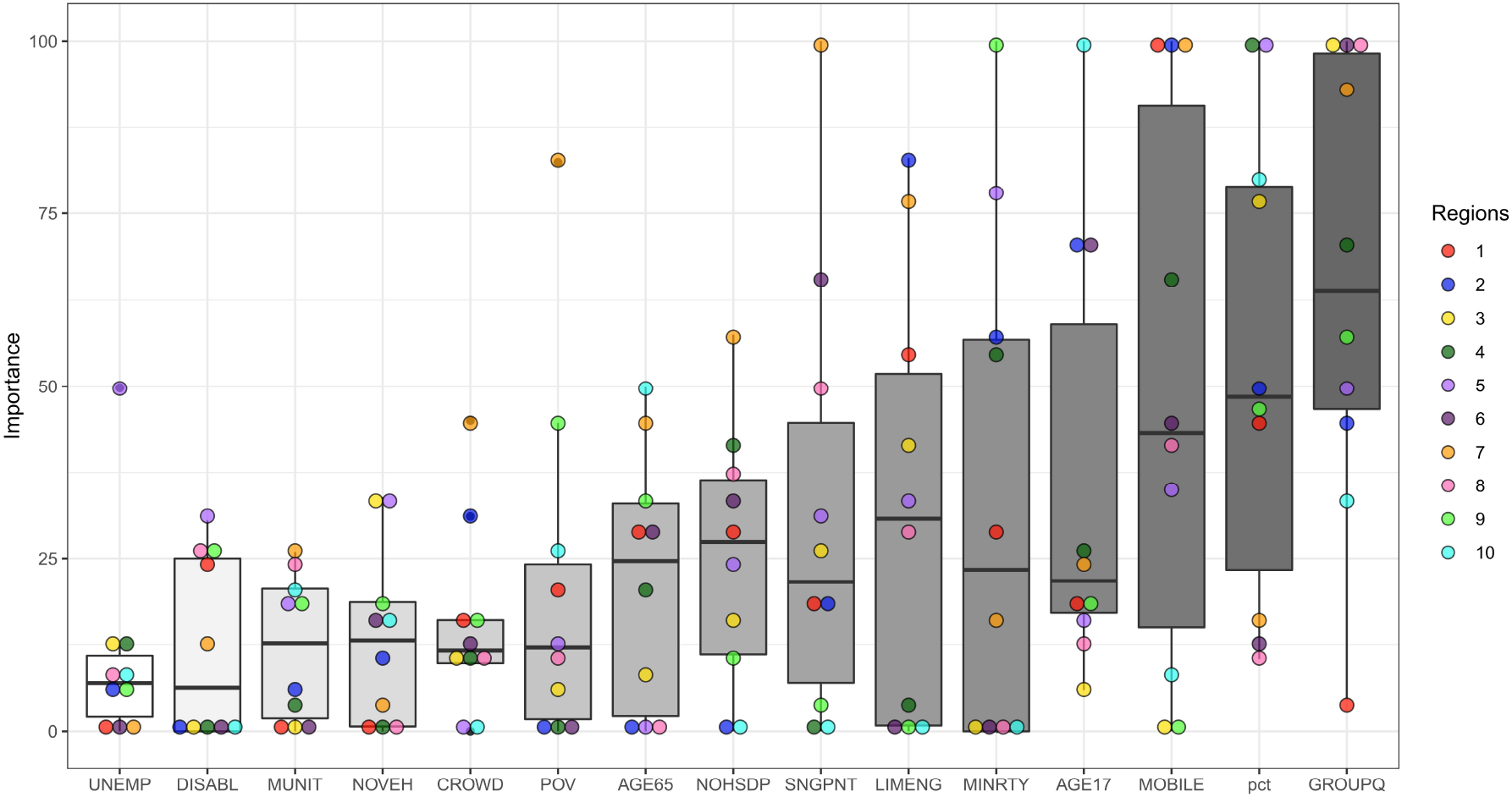
Complete pre-Delta Variable Importance Plot, organized from lowest to highest overall importance

**Figure C2.**
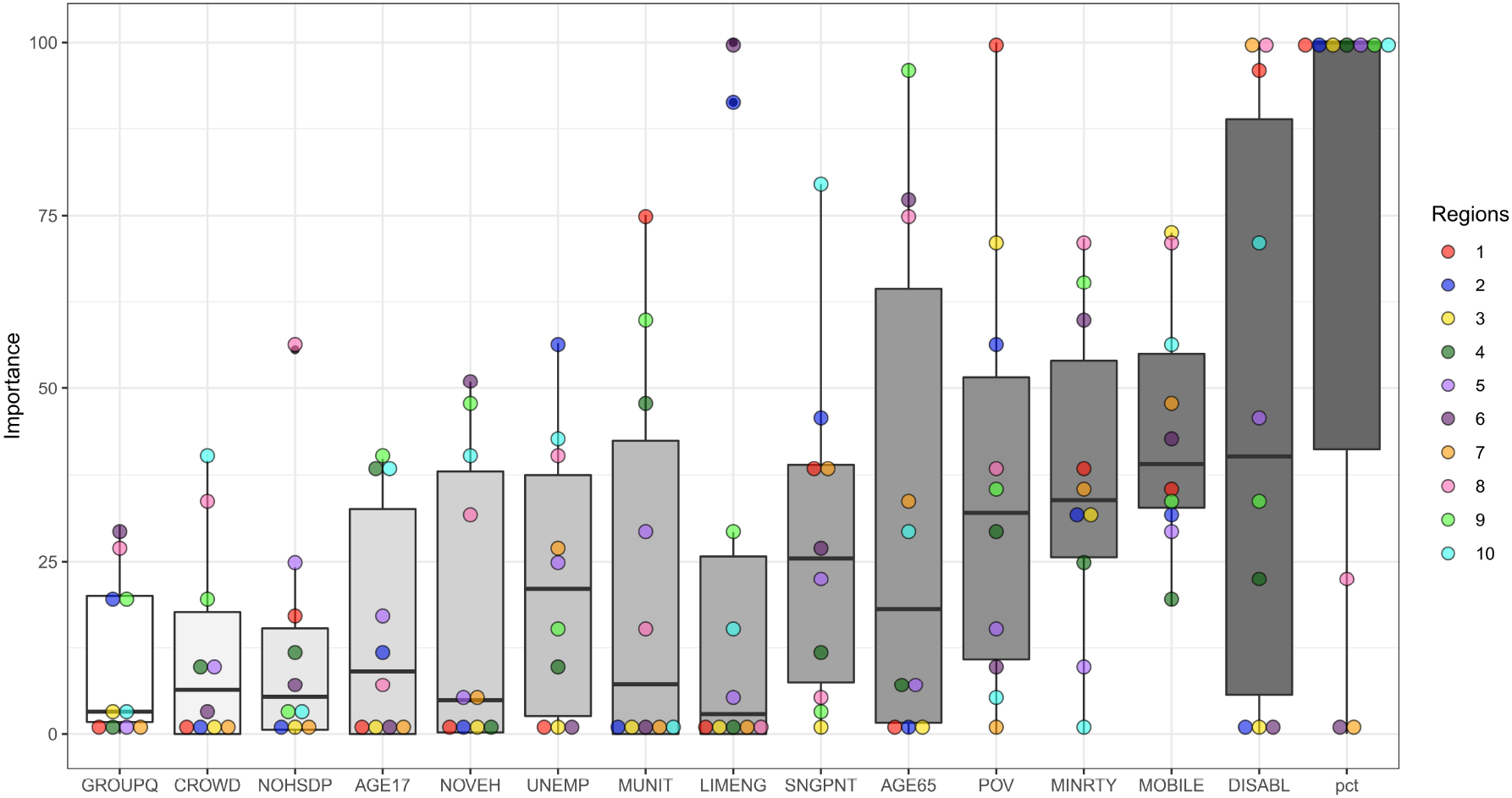
Complete Delta Variable Importance Plot, organized from lowest to highest overall importance

